# The potential clinical impact and cost-effectiveness of a variant-adapted 2024 Winter and Summer COVID-19 mRNA vaccination campaign in Australia

**DOI:** 10.1101/2024.12.18.24319245

**Authors:** Amy Lee, Michele Kohli, Michael Maschio, Keya Joshi, Ekkehard Beck, Peter Moore, Eliza Kruger

## Abstract

**Objectives:** To assess the clinical and economic impact, and cost-effectiveness, of a 2024 Winter and Summer COVID-19 Vaccination Campaign (2024 Vaccination Campaign) in Australia compared to no 2024 Vaccination Campaign.

**Design:** Modelling study using a COVID-19 susceptible-exposed-infect-recovered (SEIR) dynamic transmission model and a COVID-19 vaccination and infections consequences decision analytic model.

Setting, Participants: Australia; Winter 2024 COVID-19 vaccination targeting adults aged ≥18 years, with those aged ≥ 65 years eligible for an additional Summer 2024 dose.

Intervention: 2024 Vaccination Campaign using a Moderna variant-updated mRNA vaccine compared to no 2024 Vaccination Campaign.

Main outcome measures: Projected number of symptomatic infections, hospitalisation, deaths, and cases of long COVID prevented; quality-adjusted life-years (QALYs) gained, incremental cost of the 2024 Vaccination Campaign, and incremental cost-effectiveness ratio (ICER) of the campaign compared to No Vaccination Campaign.

**Results:** Compared to no Vaccination Campaign, a 2024 Vaccination Campaign in Australia is predicted to prevent 241,600 symptomatic infections, 13,500 COVID-19 hospitalisations, 1,200 deaths, and 11,900 cases of long COVID, representing a decrease of 16%, 23%, 26%, and 17%, respectively. This resulted in 19,200 fewer QALYs lost. COVID-19 treatment costs saved with the 2024 Vaccination Campaign was $511.7 million. This partially offset the costs associated with vaccination and adverse event treatment ($1.03 billion), resulting in an incremental 2024 Vaccination Campaign cost of $522 million for a population of 26.3 million, with 4.35 million vaccinations administered. The ICER was $27,100/QALY gained. Increasing the vaccine coverage rate to that observed with influenza vaccination prevented more cases of infection, hospitalisation, and deaths compared to the base case, with an ICER of $34,400/QALY gained.

**Conclusion:** Even in the endemic setting with high hybrid immunity, substantial clinical and economic benefits to vaccinating those aged ≥18 years against COVID-19 remain. These benefits may be amplified with increased vaccination coverage.

**Summary box:** The known: Vaccination campaigns were a cost-effective strategy to battle the clinical and economic impact of COVID-19 during the pandemic. Even in the endemic setting, COVID-19 continues to cause substantial clinical and economic burden to Australia.

The new: Even in a population with high hybrid immunity, COVID-19 vaccination campaigns continue to be cost-effective in Australia in the endemic setting. Clinical benefits are increased by improving the vaccination coverage rate, and not narrowing the targeted population.

The implications: In Australia, annual COVID-19 vaccination campaigns should continue. Increasing the COVID-19 vaccination coverage rates and including a broader recommended population is feasible with an acceptable incremental cost-effectiveness ratio.

## Introduction

Since emerging in January 2020, COVID-19 has caused substantial clinical and economic burden to Australia. In 2022-2023, there were 182,800 COVID-19 related hospitalisations, with 3.5% requiring intensive care, 1.3% requiring mechanical ventilation, and 3.6% resulting in death.^1^ In addition to the acute impact of infection, a modelling exercise estimated the long-term impacts of long COVID exceeds 102 million work hours lost in 2022, or $4.8 billion in productivity.^2^

Despite detection of SARS-CoV-2 anti-spike and anti-nucleocapsid antibodies in 99.6% and 71% of Australian adult blood donors in 2022, respectively,^3^ indicating high seroprevalence, COVID-19 continues to spread in the non-pandemic setting, driven by virus mutation, and waning of vaccine and infection-induced immunity. Thus, the Australian Technical Advisory Group on Immunisation (ATAGI) recommended vaccines for adults aged ≥75 years every 6 months. Adults aged 65-74 years and severely immunocompromised individuals aged 18-64 years are recommended to receive a vaccine every 12 months, with the consideration of an additional dose at 6 months, based on an individual risk-benefit assessment. All other adults aged ≥18 years can consider COVID-19 vaccination every 12 months.^4^ In October/November 2024, the Australian Therapeutic Goods Administration (TGA) has approved the updated JN.1 variant-adapted versions of both the Spikevax and Comirnaty vaccine.^5,6^

Despite the high COVID-19 burden, the vaccination coverage rate (VCR) in Australia remains low. As of September 11, 2024, only 19.0% of individuals aged 65-74 and 30.5% aged ≥75 years received a COVID-19 vaccine within the last 6 months. Only 6.7% aged 18-64 years were vaccinated within the last 12 months.^7^ These low rates are in stark contrast with the VCR achieved for influenza, which in 2023 in Australia was 64% for those aged ≥65 years, and 21-42% for those aged 15-64 years.^8^

Given the COVID-19 vaccination program is transitioning into the regular National Immunisation Program and reimbursement setting, it is critical to assess the economic benefits and cost-effectiveness of the vaccine. Therefore, the objective of this study is to estimate, by mathematical modelling, the cost-effectiveness of a Winter and Summer 2024 COVID-19 vaccination campaign (2024 Vaccination Campaign) in Australia using an updated Moderna variant-adapted vaccine compared to No Vaccination Campaign, over a 1-year time horizon (March 2024-February 2025). The target population is adults aged ≥18 years for the Winter dose and ≥65 years for the additional Summer dose, similar to the 2023 Winter and Summer 2024 campaigns.

## Methods

The cost-effectiveness of a 2024 COVID-19 vaccination campaign was conducted with two separate mathematical models. The projected number of vaccinations, number of infections (asymptomatic and symptomatic), protection against hospitalisations from prior vaccinations and the current campaign were calculated using a previously developed COVID-19 susceptible-exposed-infected-recovered (SEIR) dynamic transmission model. The CEA was then conducted using an infection consequences decision analytic model. Both models are described briefly herein but further details are in Kohli et al., (2023)^9^ and in the Supplementary Materials. The models were adapted for Australia using standard sources (e.g. national statistics databases and fee schedules). Epidemiological/clinical data and utility decrements inputs were identified through targeted literature reviews of databases, and manual searches of Australian government agency reports and websites. Australia-specific inputs are described below and in the Supplementary Material. All other inputs have been previously described elsewhere.^9^

## SEIR model

There are two simulation periods in the SEIR model. During the burn-in (calibration) period (January 2020-February 2024), the transmissibility of the virus over time, level of natural immunity from infections, and prior vaccination residual protection are estimated. During the second period, the 1-year analytic time horizon (March 2024-February 2025), the model is run twice; once without and once with the 2024 vaccination campaign to produce the required outcomes for each scenario.

### Winter and Summer 2024 vaccine effectiveness

As the SEIR model predicts outcomes using data from the unvaccinated population, absolute VE data are required. Although recent VE data specific to Australia are available for the original monovalent and variant-adapted bivalent vaccines, data were only available as relative VE (rVE),^10,11^ meaning the VE in those receiving the booster is expressed as additional protection gain versus those that did not receive the booster (but may have pre-existing protection from prior booster). VE for the mRNA-1273 XBB.1.5 vaccine are also available, however, they are also expressed as rVE.^12^ Therefore, real-world absolute VE against hospitalisation from the mRNA-1273 BA.4/BA.5 vaccine was used.^13^ As the study did not estimate the VE against infection, estimates from a meta-analysis^14^ of the original mRNA-1273 booster against BA.1/BA.2, were used. Monthly VE waning against infection and hospitalisation were based on a meta-analysis examining the durability of COVID-19 boosters,^15^ and applied linearly. Initial VE and monthly waning values are summarized in Table 1. As the assumed VEs are based on real-world evidence of the VE of prior vaccines against variants circulating at the time of administration, the VEs of the Winter 2024 and Summer 2024 vaccines are assumed to be the same.

**Table 1.**
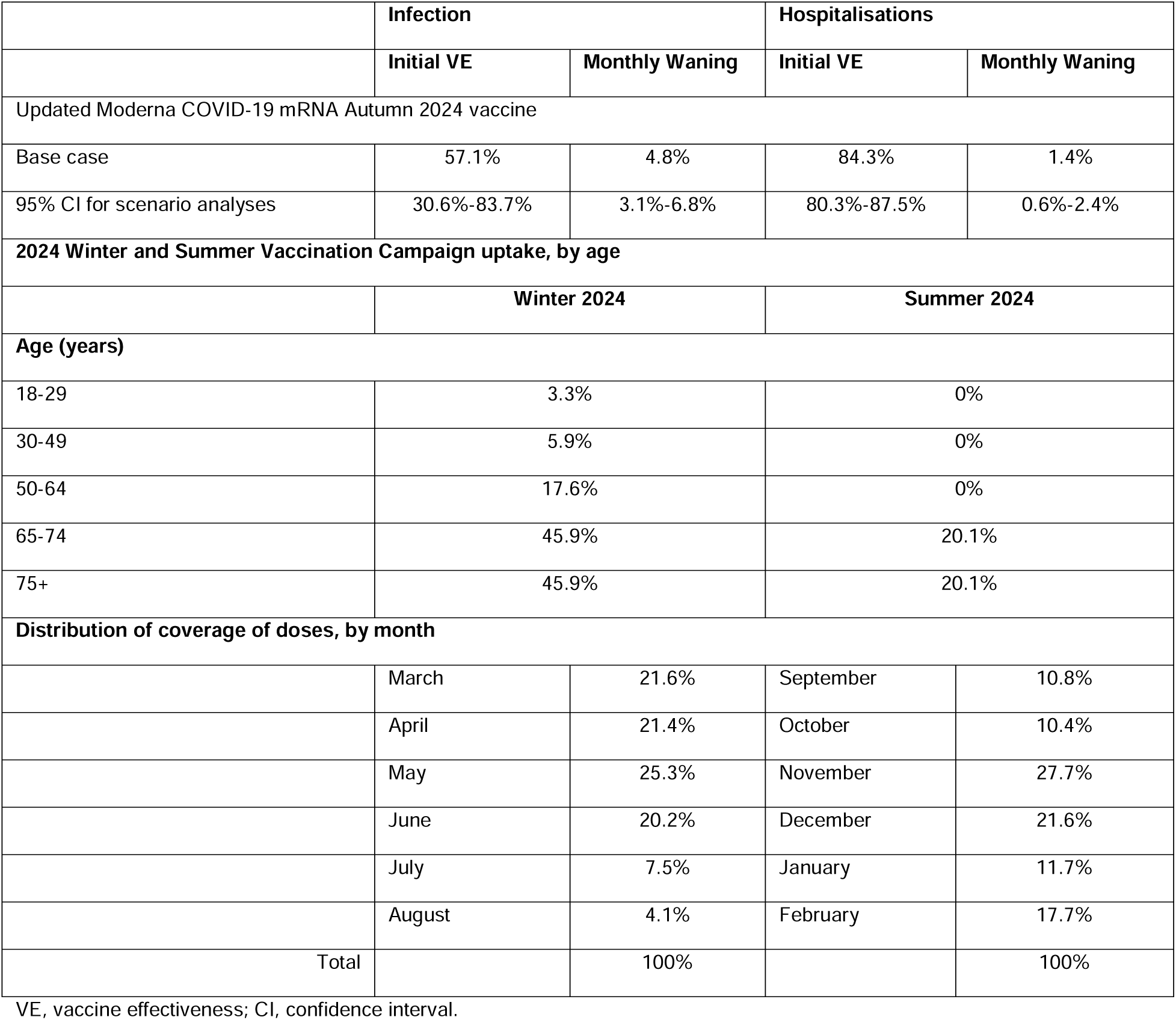
Vaccine effectiveness and coverage parameters for the Winter and Summer 2024 COVID-19 mRNA vaccines.

|  | Infection |  | Hospitalisations |  |
| --- | --- | --- | --- | --- |
|  | Initial VE | Monthly Waning | Initial VE | Monthly Waning |
| Updated Moderna COVID-19 mRNA Autumn 2024 vaccine |  |  |  |  |
| Base case | 57.1% | 4.8% | 84.3% | 1.4% |
| 95% CI for scenario analyses | 30.6%-83.7% | 3.1%-6.8% | 80.3%-87.5% | 0.6%-2.4% |
| 2024 Winter and Summer Vaccination Campaign uptake, by age |  |  |  |  |
|  | Winter 2024 |  | Summer 2024 |  |
| Age (years) |  |  |  |  |
| 18-29 | 3.3% |  | 0% |  |
| 30-49 | 5.9% |  | 0% |  |
| 50-64 | 17.6% |  | 0% |  |
| 65-74 | 45.9% |  | 20.1% |  |
| 75+ | 45.9% |  | 20.1% |  |
| Distribution of coverage of doses, by month |  |  |  |  |
|  | March | 21.6% | September | 10.8% |
|  | April | 21.4% | October | 10.4% |
|  | May | 25.3% | November | 27.7% |
|  | June | 20.2% | December | 21.6% |
|  | July | 7.5% | January | 11.7% |
|  | August | 4.1% | February | 17.7% |
| Total |  | 100% |  | 100% |

VE, vaccine effectiveness; CI, confidence interval.

### Winter and Summer 2024 vaccine coverage

In line with prior vaccination campaigns, the primary target for the Winter 2024 vaccine is the adult Australian population aged ≥18 years. The base case assumes only those aged ≥65 years can receive the Summer 2024 dose. The VCR predictions assume a winter peak (March 2024-August 2024) and a second summer peak (October 2024-February 2025) based on age-specific data from the Australian Department of Health and Aged Care from November 2022-October 2023.^16^ Total coverage and monthly proportion of total doses administered is provided in Table 1. Sensitivity analyses with assumed uptake based on the long-term average vaccination uptake for influenza were included.

## Vaccination and infection consequences model

The vaccination and infection consequences model calculates costs and QALY decrements associated with vaccination or symptomatic infection, as well as the cost-effectiveness of the 2024 Vaccination Campaign versus No 2024 Vaccination Campaign from March 2024-February 2025.

Infected patients may be asymptomatic. Only symptomatic patients are assumed to incur costs and QALY decrements, and enter into the infection consequences decision tree, where there is the risk of infection-related myocarditis. Independent of this risk, they may be hospitalised (with a risk of in-hospital mortality) due to the SARS-CoV-2 infection. Post-acute infection, there is the risk of long COVID (Figure 1). The following inputs from the SEIR model for the 2024 Vaccination and No Vaccination Campaigns were entered into the consequences model: the monthly infection incidence and residual protection at the start of the analytic time horizon. For the 2024 Vaccination Campaign only, the incremental risk reduction in hospitalisation for vaccinated individuals compared to those that did not receive the new vaccine, and number of Winter and Summer 2024 vaccinations were additionally entered. COVID-19 infection-specific clinical consequences and costs are also entered into the consequences model. These include the proportion of infections that are symptomatic, the proportion of infections that require hospitalisation in the general ward, intensive care unit (ICU), or mechanical ventilation, and readmissions, in-hospital mortality, infection-related myocarditis, and long COVID rates. Costs and QALY decrements for each event are also considered. Inputs are provided in the Supplementary Materials.

**Figure 1.**
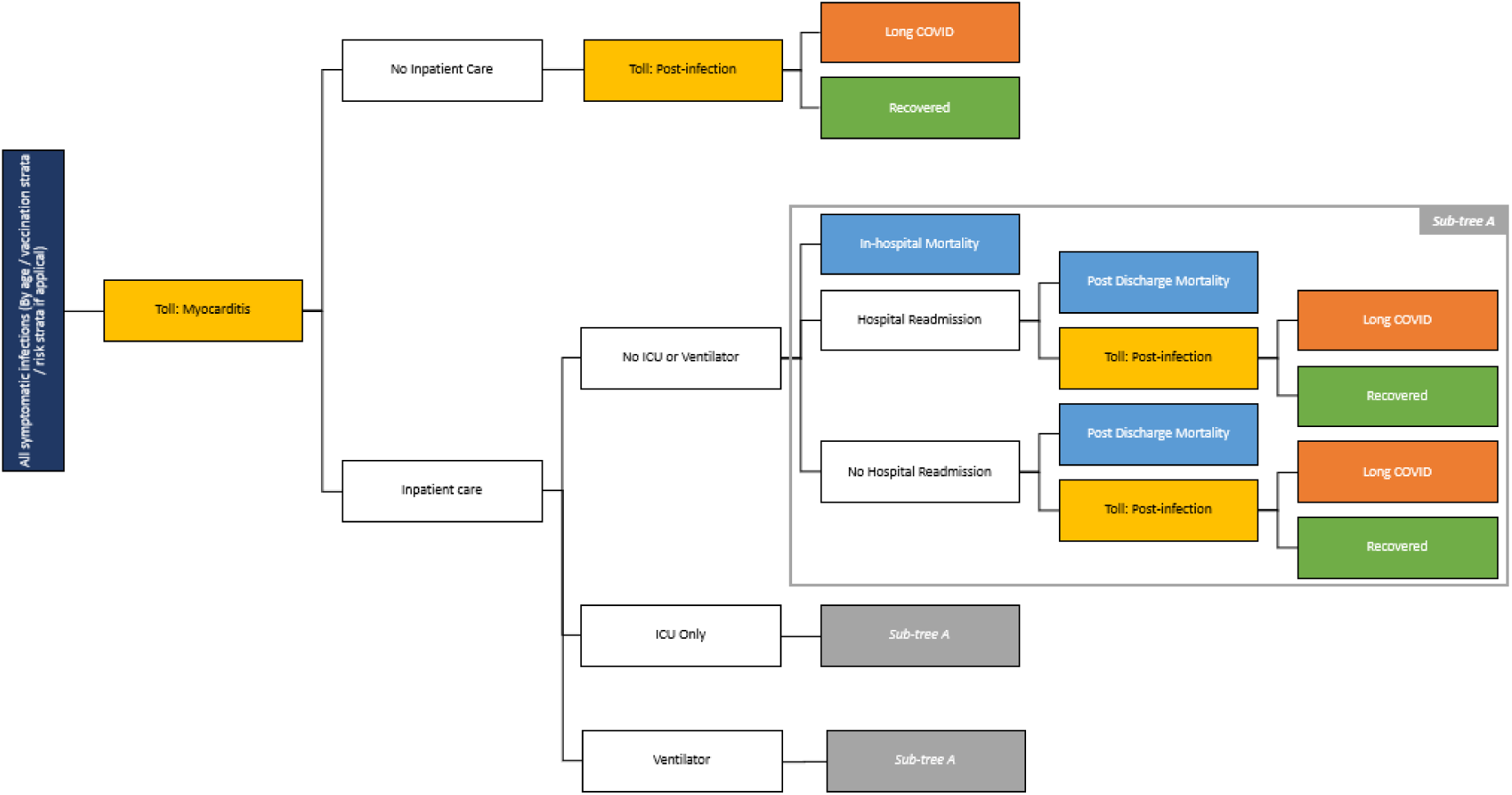
COVID-19 consequences model.

The base case analysis was conducted from the healthcare system perspective; a societal perspective sensitivity analysis includes productivity loss due to infection and vaccination. Although the analytic timeline is one year, the expected life-years and QALY decrements lost over a lifetime for a COVID-19 related death were estimated and discounted by 5% annually to present value.^17^ All other outcomes were not discounted.

The costs and consequences of vaccination, not shown in Figure 1, are also calculated in the model. Vaccination and administration unit costs, and cost and QALY decrement per treatment of vaccine-related grade 3 and above solicited local and systemic adverse events, myocarditis/pericarditis, and anaphylaxis were included. The unit cost of the updated 2024 vaccine was assumed to be $198.46 based on the USA price.^9^ An administration cost of $36.98 was added per vaccination, assuming a 1:1 ratio of administrations occurring in pharmacy versus general practitioner settings.^18,19^

Deaths were assumed to only occur in hospitalised patients as they experience more severe disease. Due to the short time horizon, all-cause mortality was excluded. Finally, it was assumed that any beneficial effect due to the use of nirmatrelvir/ritonvavir is already accounted for in the observed hospitalisation rates.

### Analyses

In addition to conducting a CEA at the population level, the number needed to vaccinate (NNV)^20^ to prevent one infection, COVID-19 related hospitalisation, death, and case of long COVID was also calculated. To assess the impact of parameter uncertainty on the robustness of the model results, deterministic sensitivity analyses (DSAs) and scenario analyses were conducted on both SEIR and consequence model parameters. Besides those already mentioned above, the SEIR model was used to generate sets of incidence scenarios by varying the transmissibility parameter (and therefore varying timing, height, or number of the infection incidence peak(s)). Residual VE due to prior vaccinations was also varied by assuming lower protection due to immune escape, and greater protection due to higher coverage before the analytic time horizon. Consequence model inputs were varied by their 95% CI where available, or other published alternate values were used, otherwise the inputs were varied by ±25%. These are presented in the Supplementary Material.

## RESULTS

Base case results are presented in Table 2. Compared to No Vaccination Campaign, the 2024 Vaccination Campaign reduces COVID-19 related hospitalisation and deaths by 23% and 26%, respectively. This is higher than the percent reduction in infections (16%) because of the additional protection of the vaccine against hospitalisations. Post-infection consequences of infection-related myocarditis and long COVID are reduced by 17%. The NNV to prevent one symptomatic infection, hospitalisation, and death is 18, 321, and 3,731, respectively. The total number of QALYs lost from the 2024 Vaccination Campaign was 19,246 fewer with the Vaccination Campaign. COVID-19 treatment cost saved with the Vaccination Campaign was $511.7 million. This partially offset the costs of vaccination-associated costs and adverse event treatments ($1.03 billion), resulting in a net vaccination campaign cost of $522 million for a population of 26.3 million ^21^ where 4.35 million vaccinations were administered. This results in an ICER of $27,126/QALY gained.

**Table 2.**
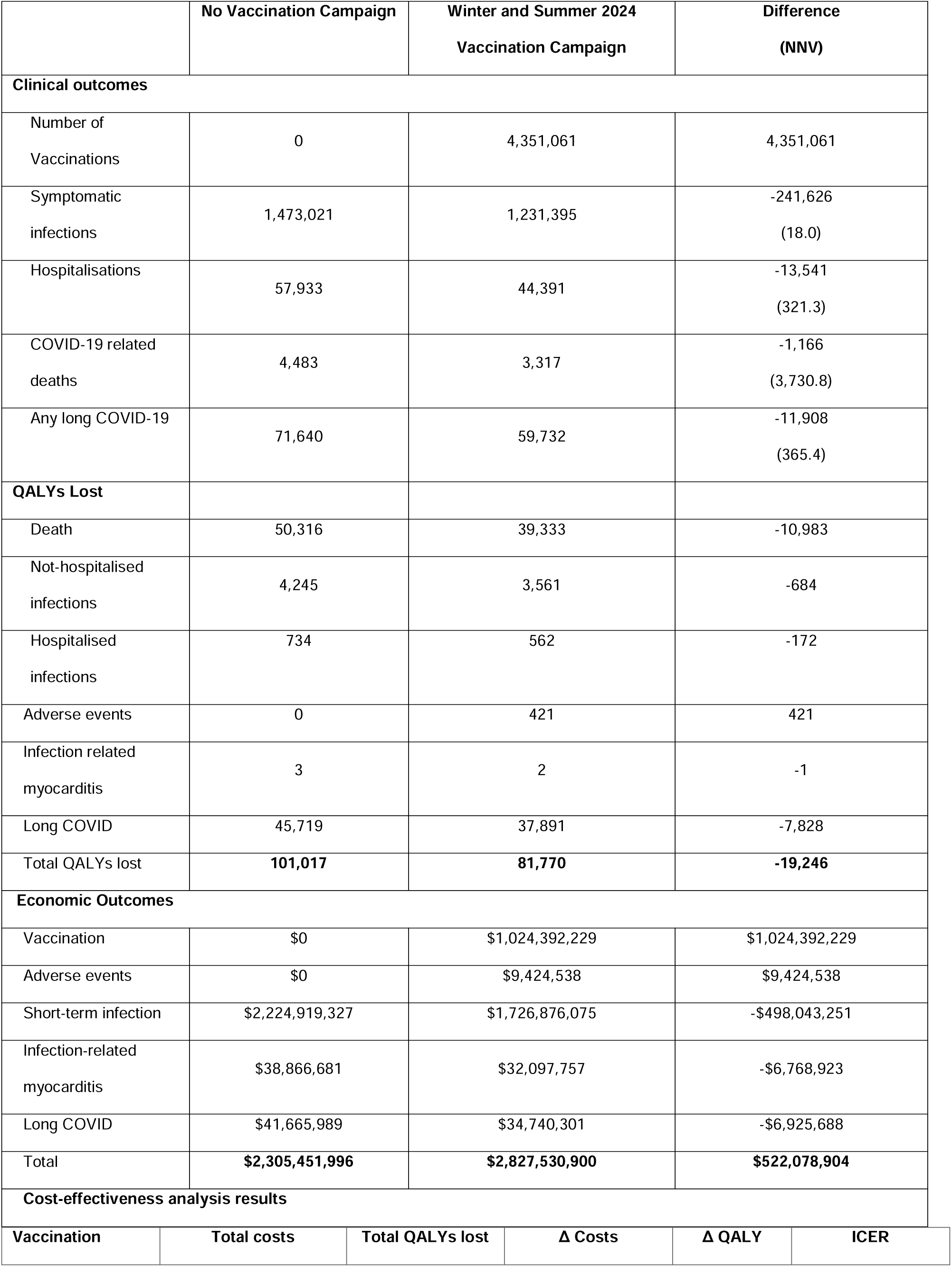
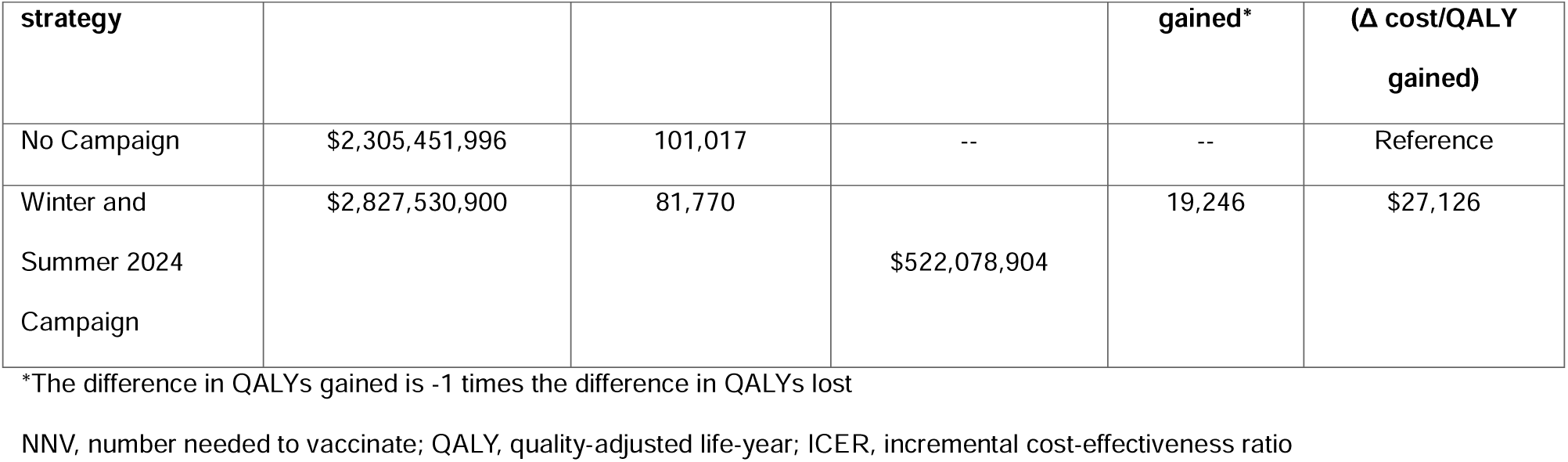
Clinical and economic base case results.

### Scenario and sensitivity analyses

Scenario and sensitivity analyses found that the ICER is highly sensitive to transmissibility parameter and incidence assumptions. All alternate transmissibility scenarios that increased infection incidence decreased the ICER. Increasing the incidence peak in May 2024 has the largest effect, decreasing the ICER to $636/QALY gained. With a higher expected infection incidence, the benefit of vaccine protection increases. Shifting the incidence peak from May 2024 to June or July 2024 also decreased the ICER as more individuals received the 2024 vaccine in the additional months before infection incidence started to increase. Decreasing the expected infection incidence, increased the ICER by ∼45% as vaccination is less impactful to a population with fewer infections to begin with. Full scenario analyses results are shown in the Supplementary Materials.

Model results were also highly sensitive to assumptions surrounding VE. SAs that decreased the VE of the Winter and Summer 2024 vaccines resulted in higher ICERs, as there was less protective effect from the vaccines. Increasing the waning rate also resulted in higher ICERs, as the duration of protection was shortened.

Limiting the vaccination campaign to those only aged 65 years and over and considering a second dose to be received in Summer increased the ICER by ∼90%, as only a small proportion of the population is vaccinated, and therefore able to benefit from direct protection. Furthermore, there is high second dose coverage in this scenario with almost half of those who received the Winter dose also receiving the Summer dose. The infection rate across the whole Australian population did not decrease to the same degree: while infections decrease by 16% in base case, the decrease was only 6% in this scenario.

Vaccinating more individuals across age groups provides more indirect protection to the population overall. Increasing the vaccination coverage to that achieved with influenza results in the prevention of more cases of infections, hospitalisation, and deaths compared to the base case while maintaining an ICER that is still cost-effective.^22^

Hospitalisation rates was a key results driver. Decreasing the rate of hospitalisations due to COVID-19 for those aged 60 years and under by 2/3 increased the ICER by ∼60%. Decreasing hospitalisation rates overall by 25% resulted in a ∼45% increase. However, even with an assumed lower rate of hospitalisation, both ICERs were still below $50,000/QALY.

Finally, assuming no post-hospitalisation mortality following discharge resulted in a 75% increase in ICER. This was an extreme analysis, as it is highly unlikely that would not be any deaths following hospital discharge.

Shifting the perspective from healthcare system only to a broader societal perspective, which includes productivity loss, the ICER decreases (improves the cost-effectiveness) by 42% to $15,765. The model predicts that vaccination will significantly enhance productivity, preventing a net $219 million in lost productivity. This is primarily due to preventing acute infections ($215 million) and mitigating severe long COVID ($9 million), and slightly offset by time loss for vaccination ($36 million) and managing adverse events ($14 million). Overall, vaccination delivers a considerable net benefit in preserving workforce productivity.

The top 15 parameters with the largest impact on ICER are displayed in Figure 2, along with the impact on results.

**Figure 2.**
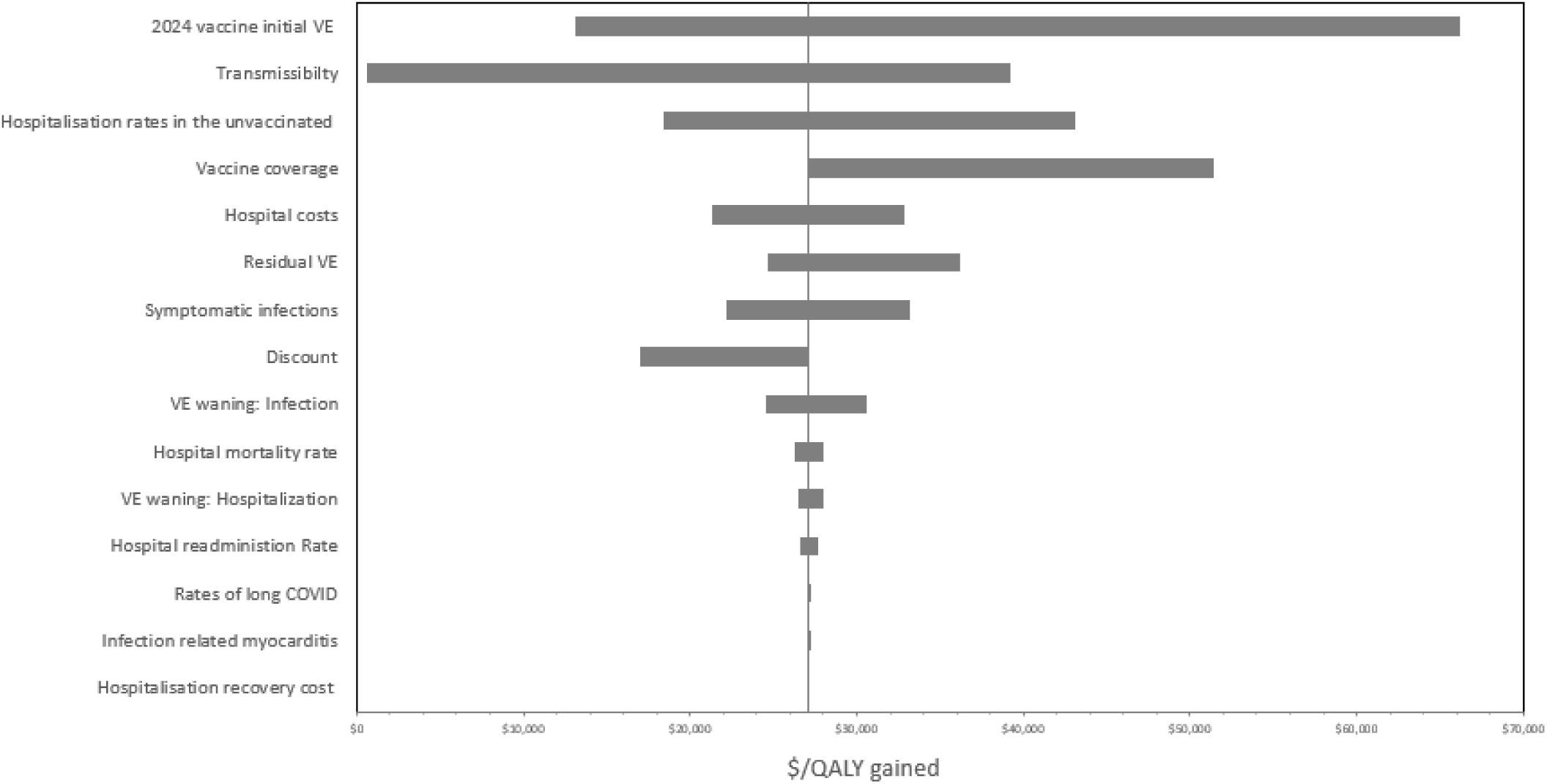
Tornado diagram: impact of the sensitivity and scenario analyses on the incremental cost per QALY gained. VE, vaccine effectiveness; QALY, quality-adjusted life-year

## Discussion

Given the significant health and economic burden posed by SARS-CoV-2 virus and the on-going uncertainty around its evolution, effective prevention strategies are essential. This analysis highlights the substantial clinical and economic benefits of a Winter and Summer 2024 vaccination campaign in Australia using an updated, variant-adapted vaccine. The campaign is predicted to be strongly cost-effective and highly impactful, potentially preventing 241,600 symptomatic infections, 13,500 COVID-19 related hospitalisations (including 680 ICU admissions), 1,200 deaths, and loss of 19,200 QALYS. These outcomes can be avoided through administration of 4.35 million vaccinations at a total net cost of $522.08 million. The analysis predicts an ICER of $27,126/QALY gained, below traditional Australian cost-effectiveness thresholds.^22^ By focusing on preventing these severe health outcomes, the campaign demonstrates strong cost-effectiveness, further emphasising the public health value of widespread vaccination.

The SEIR model accounted for protection from prior infections and vaccinations and thus allowed for a robust projection of COVID-19 infection incidence and vaccination impact for the 2024 season in a hybrid immunity setting. Even with the current high level of immunity in the population due to prior infection, vaccination or both, a Winter and Summer 2024 Vaccination Campaign continues to add protection with good value for money. This is consistent with analyses conducted in other countries (United States, United Kingdom, Japan, and Germany).^9,23-25^

Currently, the COVID-19 VCR is significantly lower than that achieved with the influenza vaccine in Australia, possibly due in part to vaccination recommendations. Unlike COVID-19 vaccines, which ATAGI recommends can be “considered” for individuals aged 18-64, the influenza vaccine is “recommended” for all individuals aged 6 months and older.^26^ This vaccination recommendation is misaligned with the observed burden from both diseases, as the clinical and economic burden of COVID-19 is larger than that of influenza.^27,28^ The scenario using VCRs similar to 2023 influenza VCRs continues to show cost-effective results. Similar to the influenza vaccine, emphasis should be placed on the importance of annual vaccination, and not total number of COVID-19 ever received.

As with all modelling exercises, there are limitations to the study. There is high uncertainty surrounding evolving variant strains and the VE of each updated variant-adapted vaccine. However, values used were based on real-world data of prior versions against evolving variants at the time of administration, much like with the application of the updated vaccine. Additionally, sensitivity analyses were conducted using the 95% CIs of the VE and monthly waning rates.

The impact of uncertainty surrounding the predicted infection incidence was tested through multiple scenario analyses where timing and size of the infection peaks were varied. All scenarios continued to yield cost-effective results.

As some Australia-specific data were missing, assumptions had to be made to estimate hospitalisation rates of symptomatic, unvaccinated SARS-CoV-2 individuals, introducing a large margin of uncertainty. Despite sensitivity analyses where these rates were reduced by 25% for the whole population, and by as much as 33% for those under aged 60 years, the ICERs still remained below ∼$43,000/QALY gained. Therefore, when extremely conservative hospitalisation rates are used, the benefits of a vaccination campaign remain.

Although there is evidence showing that long COVID has a higher impact on younger individuals, due to the lack of quantitative data, in the model, long COVID is assumed to affect all age groups equally.^29^ A conservative approach was currently taken, and the full benefit of vaccination is likely underestimated, especially from the societal perspective. However, given the observed magnitude of effect, the importance of the disease burden from long COVID even in a hybrid immunity setting, needs to be included by policy decision makers when considering the impact of vaccination.

The model did not account for a difference in outcomes for at-risk individuals (e.g. the immunocompromised). They were assumed to have the same probability of hospitalisation and death once infected as the general population. However, evidence shows more severe disease and deaths in the at-risk population. Therefore, the model may be underestimating the benefits of vaccination.

In conclusion, this study has found, that in Australia, a population with high hybrid immunity, there is still substantial clinical and economic benefits to vaccinating the adult ≥18 population against COVID-19. These benefits may be amplified with increased vaccination coverage.

## Data Availability

All data produced in the present work are contained in the manuscript

