## Supplementary material for "The potential clinical impact and cost-effectiveness of a variant-adapted 2024 Winter and Summer COVID-19 mRNA vaccination campaign in Australia"

### 1. SEIR MODEL

A previously developed Susceptible-Exposed-Infectious-Recovered (SEIR) dynamic transmission model[1] was used to project the SARS-CoV-2 infection incidence, hospitalisations, residual vaccine effectiveness (VE) from prior COVID-19 vaccinations, and number of vaccinations received, from March 2024-February 2025. The model was adapted to incorporate Australia-specific population, infection incidence and hospitalisations history, number and timing of prior vaccinations, and vaccine effectiveness (VE). The full description of the original model and explanation of movement through the model is presented in a prior publication. Here, the Australia-specific model structure, inputs, and details that differed from the original publication are provided.

The simulation with the SEIR model is run across two different periods of time. The first period is referred to as the "burn-in" or "calibration period" (January 31, 2020-Feb 29, 2024) and the second period the "analytic time horizon" (March 1, 2024-February 28, 2025).

The main structural adaptation for the Australian setting compared to the model described in the US analysis publication[1] was the change in the time horizon considered during the burn-in period. For the recent US analysis, the burn-in period was identified as the time from start of the pandemic to August 2023. For the Australian analysis, the burn-in period was extended to February 2024. This allowed evaluation of an analytic time horizon from March 1, 2024 to February 28, 2025. In addition, instead of one round of vaccination during the analytic time horizon, the Australian model includes a Winter dose to be administered starting in March 2024 and a Summer dose to be administered starting in September 2024. To accommodate these changes in the time horizon of the simulation, two additional vaccine strata were added to the model (Figure 1).

**Figure 1. Australia-specific COVID-19 SEIR model structure**

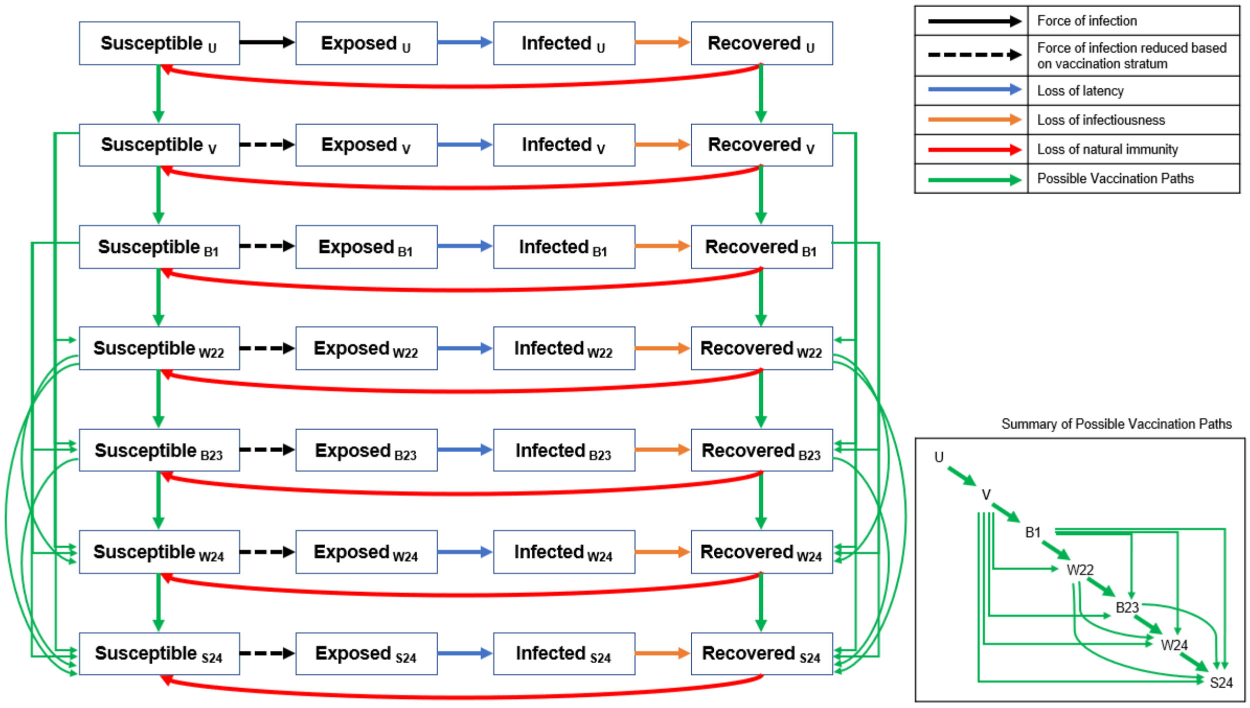

U, Unvaccinated; V, Primary Series vaccination; B1, First booster; W22, Winter 2022 booster; B23, 2023 booster; W24, Winter 2024 vaccine; S24, Summer 2024 vaccine
Figure description: Black arrows represent the movement between the susceptible and exposed compartments which is driven by the force of infection. The dashed black arrows indicate that the force of infection is modified by vaccination compared to the same transition in the unvaccinated stratum. Each vaccine stratum is associated with a unique vaccine effectiveness estimate. The blue arrows represent the loss of latency which means the infection becomes transmissible. The orange arrows represent the loss of infectiousness which means the infection is cleared and natural immunity develops. The red arrows represent the loss of natural immunity following infection and transition back to the susceptible state. Loss of latency, loss of infectiousness and loss of natural immunity is the same for all vaccine strata but can change over time. The green arrows represent the possible vaccination points. The inset box is a stylized version that shows the possible vaccination points more clearly.

The transitions between the vaccine strata during the burn-in period are governed by the historical vaccination coverage patterns. The allowed transitions that are depicted in Figure 1 with green arrows have been modified slightly from the US model because the pattern of vaccination in Australia differed from that of the US. Briefly, the timing of B2 or the Winter dose overlaps with the timing of B1 and both of these doses were the monovalent vaccine. Therefore, in the Australian model, it is possible to move directly from V (primary series completed) to either B1 or B2.

#### 1.1. Dynamic model inputs: burn-in period (January 2020 to February 2024)

In order to estimate the number of infections and existing protection to the population due to prior vaccinations (residual VE) and natural immunity at the start of the analytical time horizon, a burn-in period was used to simulate the Australian cohort from the start of the pandemic up to the end of February 2024.

Assumptions were made regarding the dates that past variants arose. According to variant data, Omicron BA.1 and BA.2 accounted for over 90% of the infections in Australia during the weeks of January 3 to 17, 2022.[2] Therefore, it was assumed that the pre-Omicron period lasted from the start of the pandemic to January 10, 2022. Omicron BA.4 and BA.5 accounted for over 90% of infections in Australia during the weeks of August 1 to August 15, 2022. Therefore, it was assumed that the Omicron BA.4/5 period began on August 8, 2022.

Within the model, changes were made to VE and to the waning rate of natural immunity according to the dates as described in the sections below. Essentially, the residual VE from previous vaccines and the proportion of the population in the R states drops at the start of a new variant period to reflect the impact of immune escape. While variants emerge gradually over time, an assumption regarding the date of variant change is required because the model cannot accommodate gradual change in vaccine and natural immunity over time. The current approach is conservative, as it increases historical vaccine and natural immunity and decreases the number of infections observed in projections.

#### 1.1.1. Number of susceptibles

All individuals in Australia were included in the model and are considered susceptible to COVID-19 infection at the start of the simulation (January 31, 2020). The size of the Australian population in 2022 by age was obtained from the Australian Bureau of Statistics.[3] The simulation used a closed cohort which did not consider births, deaths, immigration or emigration and therefore the 2022 population was considered to be most appropriate.

#### 1.1.2. Mixing patterns / contact matrices

The age-specific mixing patterns are based on country-specific contact matrices. As behaviours that impacted contact changed during the pandemic, the matrices are modified by a mobility index that accounts for social distancing and mask use.

Data on the age-specific mixing patterns in the general population for Australia were obtained from Prem and colleagues.[4] In the published contact matrices, the population was partitioned into 5-year age bands, and all individuals aged 75 years and older were grouped together. For this analysis, the age-specific mixing patterns were first converted into 10-year age bands. Then, we assumed symmetry between the age groups (i.e., an effective contact between someone in age group *i* with someone from age group *j* is the same as an effective contact between someone in age group *j* with someone from age group *i),* weighted by the population estimates in age groups *i* and *j*.

Thus, $c_{ij} = \frac{1}{N_j} \times \frac{(c_{ij}^{*} \times N_j) + (c_{ji}^{*} \times N_i)}{2}$ ,

where:

- $c_{ij}$ is the number of effective contacts between someone in age group *i* with someone from age group *j*
- $c_{ij}^{*}$ is the number of effective contacts between someone in age group *i* with someone from age group *j* based on the fact that the original age-specific mixing patterns were first converted into 10-year age bands
- $N_i$ is the population size in age group *i*.

During the pandemic, the rate of contact was reduced by behaviours such as social distancing and mask use. The magnitude of the impact is estimated as described in the next section.

##### 1.1.3. Social distancing and mask use: overall scaling factor

From January 31 2020 to June 24, 2022, it was assumed that regular patterns of interaction were modified because of social distancing and use of masks to reduce effective contacts between individuals. Data on social distancing patterns and mask use were obtained from the IHME and used to adjust the base contact matrix based on these factors.[5] Daily estimates on social distancing patterns and mask use were obtained from the IHME for the time period February 2020 through June 24, 2022, inclusive. For the time period after July 31, 2022, a single seasonality parameter was assumed to replace the social distancing patterns and mask use data. Between these two time points (June 24, 2022 and July 31, 2022), a linear interpolation between the scaling factors estimates was assumed to avoid an abrupt transition between these two methods.

For the base case, it was assumed that mask use returned to pre-pandemic levels (0%) from July 31, 2022 onwards. We also assumed that social mobility would return to normal (baseline) after this date and stay at this level for the remaining time period.

Daily estimates of mask use were obtained from IHME.[5] Mask use represents the percentage of the population who say they always wear a mask in public. It was assumed that 100% mask usage is associated with a 30% reduction in transmission and that a reduction in mask usage impacts the reduction in transmission proportionately.[6]

Daily estimates of the change in mobility were obtained from IHME[5] and was applied to reduce the number of contacts per person. Since age-specific data on changes in mobility were not available, the impact was applied equally to all age groups. After July 31, 2022, a standard sinusoidal function was applied to vary the rate of contact by season.[7] The assumed peak was August 15 to account for greater time spent indoors in the winter. The assumed trough was February 15 to account for greater time spent outdoors in summer. The trough and peak were 95% and 105% of normal contact respectively.[8]

##### 1.1.4. Historical vaccination coverage

The weekly age-specific data on vaccination coverage for April 2021 to September 2023 were obtained from COVID-19 vaccine rollout reports and data from the Australian Government Department of Health and Aged Care.[9] These data were used to estimate the proportion of the population that moves into the primary series and booster model strata.

#### 1.1.5. Vaccine effectiveness inputs for the burn-in period and at the start of the analytic period

Vaccine effectiveness and waning inputs for the burn-in period and at the start of the analytic period was determined using the same method and values published in Kohli et al., with the exception of Ad26.COV02.S which isn't included for Australia, and ChAdOx1, which wasn't included in the US manuscript, and therefore is not discussed here. These were weighted by the market share of each vaccine obtained from the National Centre for Immunisation Research and Surveillance (NCIRS) AusVaxSafety system,[10] to estimate single VE values primary series and booster doses, by variant at time of administration.

#### 1.1.6. Transmissibility: February 2020 to August 2023

During the first 30 days of the burn-in period, an initial 6,226 infections was used to seed or start the pandemic. This number is equal to the number of infections estimated by the IHME over the first 30 days in Australia.[5] Model calibration was then conducted to estimate the transmissibility parameter that reflects cases of COVID-19 experienced in Australia from February 2020 through August 2023. The analytical choices for the calibration process were made considering the recommendations of Vanni and colleagues.[11] The transmissibility parameter was allowed to vary on a daily basis. For the calibration process, transmissibility parameters were manually varied. The calibration targets and the goodness-of-fit measures used are described along with the results of the calibration below.

#### 1.1.7. Calibration targets

As the collection of COVID-19 related infections data has changed over time, two different outcomes of interest, or targets, were used for the calibration process.

For the first part of the pandemic, the target was the total number of infections in the population. This includes all asymptomatic and symptomatic infections, whether or not they have been reported. While data on the number of infections are collected by public health authorities, these figures reflect the number of reported infections and not the true number of infections. There are multiple factors that lead to under-testing and under-reporting of infections including patient and physician behaviour as well as technical limitations to testing. Therefore, output from the modelling done by the IHME, which has been corrected for these biases and includes all infections (symptomatic and asymptomatic), was used as the calibration target. The IHME model was chosen because it was one of the models considered by the United States Centers for Disease Control over time in their ensemble model forecasts.[12] Prior to the pandemic, IHME had developed methods of collecting data globally, which they were able to do in real time during the pandemic in order to create projections for all countries across the globe. IHME has produced multiple publications on their COVID-19 model and the structural and input modifications over time.[13] Finally, the detailed outputs from their model are publicly available. The calibration target was the daily incidence of reported COVID-19 cases (symptomatic and asymptomatic), as reported by the IHME during the period of February 4, 2020 through October 31, 2022.[5] While IHME reported outputs after October 31, 2022, the overall reporting of infections became less reliable overall. Therefore, the source of the data for the calibration changed after this date.

For the time period November 1, 2022 through July 30, 2023, the calibration target was based on the number of cases, estimated from the COVID-19 Australia: Epidemiology Reports published in Communicable Disease Intelligence (CDI).[14,15] The case definitions are based on confirmed and/or probable case as specified in the epidemiological report. Since these definitions do not include all estimated infections, as in the IHME data, the counts of cases were adjusted. For the time period May 9, 2022 through October 31, 2022 there are data from both sources and the case definition in the epidemiological reports is consistently defined as: polymerase chain reaction confirmed and rapid antigen testing probable cases. The total number of infections estimated by IHME was divided by the number of cases reported in the epidemiological reports to estimate the adjustment factor used to inflate the case counts for the time period November 1, 2022 through July 30, 2023.

$$\frac{\#\ infections\ estimated\ by\ IHME}{\#\ cases\ reported\ in\ the\ epidemiological\ reports} = \frac{15{,}132{,}913}{3{,}452{,}327} = 4.4$$

There were no data on the number of infections or hospitalisations due to COVID-19 available after July 30, 2023. For the period of August 1, 2023 to February 29, 2024, the trends in hospitalisations over time were used to guide the calibration process.

##### 1.1.7.1. Goodness-of-fit measure

Model calibration was performed in a qualitative way by visually comparing the daily incidence of COVID-19 cases predicted by the model and the values obtained from the IHME data for the first time period (February 4, 2020 through October 31, 2022) and the adjusted CDI Epidemiology Reports case data for the second time period of the calibration, November 1, 2022 through July 30, 2023.

For the dates of August 1, 2023 to February 29, 2024, the lack of calibration target meant that there was no formal goodness-of-fit. Instead, the number of infections were calibrated so that the trends in infection over time mirror the trends in hospitalisation over time.

##### 1.1.7.2. Results of the calibration

Transmissibility parameters were manually varied, and linear interpolation was used to estimate values over time. The resulting comparison plot of the daily number of incident cases of COVID-19 predicted from the calibrated model and the corresponding estimates from the IHME and the adjusted CDI Epidemiology Reports case data is presented in Figure 2.

**Figure 2. Comparison of daily incidence of COVID-19 cases (calibrated model estimates vs. targets)**

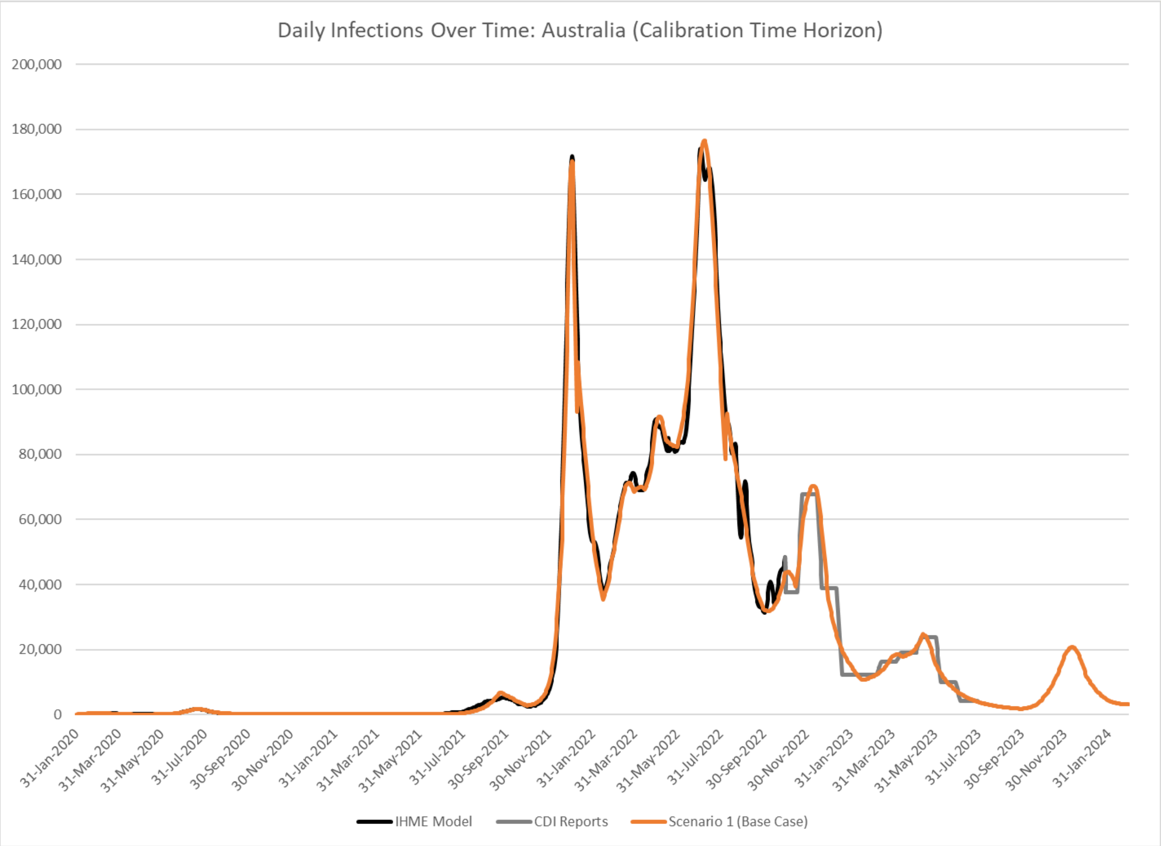

CDI, Communicable Diseases Intelligence; IHME, Institute for Health Metrics and Evaluation

##### 1.1.7.3. Validation: number of hospitalisations

The reported number of hospitalisations for the period of July 1, 2021 to June 30, 2022 were estimated for the model age groups from data from the Australian Institute of Health and Welfare.[16] Following the calibration to infections, the hospitalisation rates in the unvaccinated were derived as described in Section 2.1.2. When these hospitalisation rates are used within the model, the number of hospitalisations predicted by the model for this same time period matches the expected cases.

### 1.2. Dynamic model inputs: Analytical time period (March 2024 to February 2025)

In this section, the inputs for the dynamic model required to project incidence of COVID-19 infection without 2024 COVID-19 vaccination are described.

After February 29, 2024, the transmissibility parameters were varied during the analytic period considering the values over the last year of the calibration period to produce six different patterns:

1) Base case: the transmissibility parameter is changed so that a peak in incidence occurs in May.
2) Scenario 1: the transmissibility parameter is changed so that a peak in incidence occurs in June.
3) Scenario 2: the transmissibility parameter is changed so that a peak in incidence occurs in July.
4) Transmissibility scenario 3: May 2024 peak increased
5) Transmissibility scenario 4: May 2024 peak decreased
6) Transmissibility scenario 5: May 2024 peak; Second peak In December 2024

### 2. VACCINATION AND INFECTION CONSEQUENCES MODEL

Similar to the SEIR model, the vaccination and infection consequences model is described in detail elsewhere.[17] Australian-specific inputs and differences in the model are described in the main manuscript or in the sections below. All others are available in the previous publication by Kohli et al. (2023).[1] Australia-specific inputs are displayed in Table 2. Where further details are required, they are provided below.

#### 2.1. Vaccination and Infection Consequences Tree Inputs

##### 2.1.1. Proportion symptomatic infections

It was assumed that only SARS-CoV-2 infected symptomatic individuals incur costs and QALY decrements associated with infection. The proportion of all infections that are symptomatic, by age, was obtained from an Australian study by Gomez et al.,(2022).[18] However, as the data were not age specific, age distributions from a study by Wang et al., (2023)[19] was applied to the overall proportion reported by Gomez to estimate age-specific values for Australia.

#### 2.1.2. Hospitalisation rates in the unvaccinated

Australian data on hospitalisation rates of symptomatic, unvaccinated, SARS-CoV-2 infected individuals are unavailable. Therefore, the proportion of patients hospitalised in the population, regardless of vaccination status, was calculated as a ratio of the number patients hospitalised to the number of cases (infections). Data on number of cases was obtained from the Australian COVID-19 epidemiology reports from the Australian Government Department of Health and Aged Care.[14] Number of hospitalised cases was obtained from the Australian Institute of Health and Welfare (AIHW) admitted patients' activity data for the period of July 1, 2021 to June 20, 2022.[16] Following calibration of the SEIR model, it was used to predict the number of hospitalisations by age group for this time period was determined. The rates were adjusted by age group by a multiplication factor so that the predicted number of hospitalisations were similar to the observed number of hospitalisations. For most age groups, the hospitalisation rates needed to be adjusted upwards, which is expected as the vaccine does have an impact on the rate of hospitalisation given infection. In the younger age groups, the rates needed to be adjusted downwards. This is likely because vaccine coverage rates in these age groups are lower and the number of infections used for the denominator of the hospitalisation rate in the population is more highly under-reported than in the older age groups (Table 1).

**Table 1. Hospitalisation rate in the population and adjusted hospitalisation in the unvaccinated predicted during the model for the period of July 1, 2021-June 30, 2022.**

| Age group | Hospitalisation rate (population) | Hospitalisation rate (unvaccinated) |
|---|---|---|
| 0-14 years | 3.57% | 2.14% |
| 15-19 years | 1.98% | 1.78% |
| 20-29 years | 2.12% | 3.18% |
| 30-39 years | 2.50% | 3.25% |
| 40-49 years | 2.54% | 3.56% |
| 50-59 years | 3.48% | 4.52% |
| 60-64 years | 5.08% | 9.65% |
| 65-74 years | 8.25% | 15.68% |
| 75+ years | 20.19% | 42.40% |

#### 2.1.3. Level of hospital care and readmission rates

The distribution of hospitalised patients based on level of care received was derived using the number of patients across general ward, ICU, and ventilation published by AIHW patients' activity data.[16] The age categories provided in the data sources are not the same as those required by the model. Hence, the data were weighted and adjusted across the age bands using the total Australian population distribution.[3] Table 2displays the proportion of hospitalisation along with the distribution by level of care by various age cohorts considered in the model.

The proportion of patients who were subsequently admitted to hospital (i.e., readmission) was obtained from a global SLR conducted by Ramzi et al. (2022) which reported 1-year hospital readmission rates at 10.68% in patients aged <=65 years, and 12.83% and 15.28% for patients aged 65-74 years and >=75 years, respectively.[20] These findings align with those derived from a study conducted by Hodgson et al. (2021) in the Australia setting (time period: March to October 2020).[21] This Australian prospective observational study reported that 10.7% of patients were readmitted into hospital within 6-months of discharge from hospital. It was assumed that the readmission rate is same across the age bands, due to lack of published data by age-group. The data reported by Hodgson et al. (2021) were chosen for use in the base case as they are specific to the readmission rate in an Australian context.

##### 2.1.4. Long COVID

As per the World Health Organization (WHO), post COVID-19 condition, also known as long COVID, occurs in individuals with a history of probable or confirmed SARS-CoV-2 infection, usually 3-months from the onset of COVID-19. Symptoms last for at least 2 months and cannot be explained by an alternative diagnosis.[22]

There is much about long COVID that is unknown and there is a scarcity of published information globally. This is the case for Australian-specific data concerning the prevalence and impact of long COVID. A cohort study conducted by Liu et al., (2021) on people who were infected early in the pandemic (infections confirmed between January and May 2020) in New South Wales followed the infected people until three months post-infection. The study estimated that around 5% of total survivors and 9.6% of hospitalised patients still had symptoms at three months post-infection.[23] These findings align with those reported by the Australian National University Centre for Social Research and Methods who estimated long COVID prevalence to be 4.7% based on a survey (n=3,510) conducted between 8$^{th}$ to 22$^{nd}$ October 2022.[24,25] Additionally, a survey conducted by Woldegiorgis et al. (2023) in Australia reported that 18.2% (n=2,130) of respondents met the case definition for long COVID during the Omicron period.[26] The figures provided by Liu et al. (2021) were based on NSW data from January to May 2020, whereas the other two sources reported estimates based on 2022 survey data (i.e., post-Omicron). To be conservative, we assumed a 4.7% prevalence rate for long COVID for non-hospitalised patients based on the Australian National University Centre results.[25] This is consistent with other Australian studies.[23,27] Furthermore, we assumed the rate of 9.6% for hospitalised patients.[23]

### 2.2. Costs and QALY decrements

Costs included in the vaccination and infection consequences model are provided in Table 2. QALY decrements are provided in the prior publication.[1] Where further details are required to explain how values are derived, details are provided below.

#### 2.2.1. Hospitalisations

A retrospective cohort study conducted by Markey et al. (2023) reported daily costs associated with admission into general ward, ICU, and requiring ventilation.[28] This was used as the base case for the model.

Markey et al. (2023) reported the proportion of ventilation-related expenses is 25.80% in the overall costs of ICU + ventilation.[28] Using this estimate, the daily ICU cost was derived by subtracting the ventilation-associated costs from overall costs of ICU + ventilation. These daily costs were multiplied with LOS reported by latest Australian COVID-19 epidemiological report to derive cost per hospitalisation event.[14]

#### 2.2.2. Adverse event costs

The model included grade 3 local and systemic adverse events (AEs). Local events are injection site pain, axillary swelling/tenderness, swelling (hardness) or erythema (redness). Systemic events are fatigue, headache, myalgia, arthralgia, chills, nausea/vomiting, and fever. In addition, costs associated with anaphylaxis and vaccination-related myocarditis were included.

AusVaxSafety is a national, active COVID-19 vaccine safety surveillance program in Australia. They report that 0.03% of patients receiving Moderna BA.1 or Moderna BA.4/5 report going to the doctor or emergency department in the days after receiving vaccination.[10] AusVaxSafety does not ask the reason for the visit, therefore medical attendance may or may not be related to any adverse events. Medical resource utilisation for different adverse events was not available in Australia. Therefore, we use the medical attendance reported by AusVaxSafety to infer medical resource attendance by adverse event. We assume that all cases of anaphylaxis and myocarditis require medical resource utilisation and that the same proportion of grade 3 events require medical attendance and solve the proportion to sum to 0.03% (grade 3 local, 0.097%; grade 3 systemic 0.151%; anaphylaxis 0.05%; myocarditis 0.002%). There were no grade 4 adverse events

#### 2.2.3. Indirect costs

Indirect costs were included in the societal perspective sensitivity analysis. These values are described below.

Estimation of indirect costs requires three different type of inputs- proportion of population in the work force, average wage, and time lost due to infection (Table 2) Data on population in work force and average wage were acquired from the Australian Institute of Health and Welfare,[29] and the Australian Bureau of Statistics,[30] respectively.

For time loss due to vaccination data were leveraged from AusVaxSafety.[31] 8% of individuals reported missing work, study, school or routine activities after receiving Moderna BA.4/5 vaccination. The majority reported missing 1 day or less. To be conservative, we assume that 8% missed an entire day, therefore on average 0.08 days were lost following vaccination.

No Australian studies reported the time loss associated with COVID-19 infection, hence data from US studies were used to estimate productivity losses for symptomatic infections (for not hospitalised and hospitalised patients) and post-hospitalisation.

An Australian study conducted by Woldegiorgis et al. 2022 reported the proportion of long COVID patients whose employment hours were affected.[26] Time lost due severe long COVID were obtained from a US study, Ham et al. (2022).[32] All indirect cost inputs are available in Table 2. Where reported age groups differed from age groups in the model, values were weighted by population size to calculate inputs need for the model.

#### Table 2. Clinical and economic inputs

| Model Parameter | Value |
|---|---|
| **Infection consequences clinical inputs** | |
| Proportion symptomatic, by age[19,33] | |
| 0-14 years | 60.60% |
| 15-19 years | 58.42% |
| 20-29 years | 63.55% |
| 30-39 years | 70.28% |
| 40-49 years | 72.64% |
| 50-59 years | 74.39% |
| 60-64 years | 77.14% |
| 65-74 years | 80.98% |
| ≥75 years | 86.73% |

| | | | |
|---|---|---|---|
| Proportion seeking outpatient care (%)[34] | 51.5 | | |
| # of outpatient visits per patient[34] | 20.5 | | |
| # of emergency department visits per patient[35] | 0.05 | | |
| Omicron-adjusted hospitalisation locations of care, by age (%)[16] | | | |
| | No ICU or ventilation | ICU | ICU with ventilation |
| 0-14 years | 99.09% | 0.68% | 0.23% |
| 15-19 years | 97.98% | 1.43% | 0.59% |
| 20-29 years | 97.59% | 1.70% | 0.71% |
| 30-39 years | 96.74% | 2.26% | 1.00% |
| 40-49 years | 94.60% | 3.61% | 1.79% |
| 50-64 years | 92.25% | 5.16% | 2.59% |
| 65-74 years | 91.50% | 5.72% | 2.79% |
| ≥75 years | 91.79% | 5.59% | 2.63% |
| Hospital readmission rates, by initial hospitalisation location of care, all ages (%):[20] 10.7% | | | |
| In-hospital mortality rates, by age and location of care(%)[16] | | | |
| 0-14 years | 0.03% | | |
| 15-19 years | 0.09% | | |
| 20-29 years | 0.10% | | |
| 30-39 years | 0.15% | | |
| 40-49 years | 0.50% | | |
| 50-59 years | 1.19% | | |
| 60-64 years | 1.57% | | |
| 65-74 years | 3.32% | | |
| ≥75 years | 6.95% | | |
| Post-hospitalisation mortality rates (within 60 days of discharge), all ages[36,d] | | | |
| No ICU or ventilation (general ward) | 5.4% | | |
| ICU with/without ventilation | 10.4% | | |
| Long covid rates, by location of care in acute period, all ages | | | |
| No ICU or ventilation (general ward)[25] | 4.7% | | |
| ICU with/without ventilation[23] | 9.6% | | |
| **Infection consequences economic inputs** | | | |

| | |
|---|---|
| Physician visit[37] | $188.79 |
| Emergency department visit[38] | $887.92 |
| Hospitalisation, general ward[28,39,40] | $28,692.72 |
| Hospitalisation, ICU[28,39,40] | $42,749.66 |
| Hospitalisation, mechanical ventilation[28,39,40] | $65,594.69 |
| Hospitalisation recovery cost[37] | $239.01 |
| Infection-related myocarditis | $25,105.29 |
| Long covid[26,28,37-40] | $581.60 |
| **Vaccination consequences clinical inputs** | |
| Vaccine-related myocarditis/pericarditis[41] | |
| 20-29 years | 4.6/100,000 |
| 30-39 years | 1.8/100,000 |
| Vaccination consequences economic inputs | |
| Vaccine, per unit[1] | $198.46 |
| Vaccination administration[42,43] | $31.84 |
| Adverse events | |
| Grade 3 local[43,44a] | $30.83 |
| Grade 3 systemic[43,44b] | $8.13 |
| Anaphylaxis[38,39,45] | $975.87 |
| Vaccination-related myocarditis[38,39,45] | $12,373.83 |
| **Indirect cost inputs (only used for societal perspective)** | |
| Proportion in work force[29] | |
| 0-14 years | 0.0% |
| 15-19 years | 69.3% |
| 20-29 years | 78.0% |
| 30-39 years | 86.2% |
| 40-49 years | 85.5% |
| 50-59 years | 76.6% |
| 60-64 years | 67.9% |
| 65-74 years | 14.8% |
| 75+ years | 14.8% |
| Average wage per week [30] | $1,394.10 |
| Time loss (days) | |
| Vaccination[31] | 0.08 |

| | |
|---|---|
| Symptomatic infection, not hospitalised[46] | 3.57 |
| Inpatient care, general ward[16] | 10.90 |
| Inpatient care, ICU[16] | 14.30 |
| Inpatient care, ventilator[16] | 18.40 |
| Post-hospitalisation[36] | 33.43 |
| Severe long COVID[32] | 24.66 |
| Of those with long covid, $ reporting complications affected employment hours[26c] | |
| 0-14 years | 0.0% |
| 15-19 years | 21.20% |
| 20-29 years | 21.20% |
| 30-39 years | 16.30% |
| 40-49 years | 19.00% |
| 50-59 years | 14.60% |
| 60-64 years | 16.4% |
| 65-74 years | 23.5% |
| 75+ years | 31.70% |

[a]1.98% of patients experiencing AE required a physician visit; 100% of patients required a steroid cream
[b]1.98% of patients experience AE required a physician visit and paracetamol.
[c]individuals who lost time from work were considered to have severe long COVID. In the infection consequences models, these values were therefore used as inputs for the proportion who have severe long COVID.

### 2.3. Additional Results

The number of symptomatic infections, per month and age group in both the 2024 Vaccination Campaign and No Vaccination Campaign, by age, and month, are displayed below (Figure 3). It is clear that in all age groups, the 2024 Vaccination Campaign prevented infections over time.

**Figure 3. Number of symptomatic infections, by age group and month, with the 2024 Vaccination Campaign, and with No Vaccination Campaign (Month 1 = March 2024)**

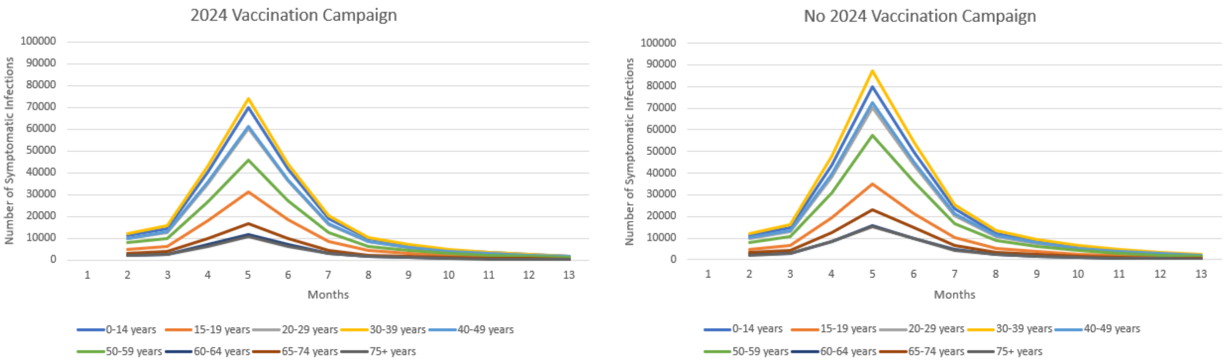

### 2.4. Sensitivity analyses

Table 3 provides the list of sensitivity/scenario parameters and values tested. The top 15 parameters with the largest impact on ICER are displayed as a tornado diagram in the main manuscript.

### Table 3. Sensitivity and scenario analyses results

| Variable or assumption | Base-case value | Plausible alternative(s) or range of values | Incremental cost / QALY gained | Description of impact on ICER |
|---|---|---|---|---|
| Base Case | | | $27,126 | |
| Transmissibility: | Peak in May 2024 | Scenario 1: Peak in June 2024 | $18,020 | -33.57% |
| Transmissibility: Scenario 2 | Peak in May 2024 | Scenario 2: Peak in July 2024 | $8,843 | -67.40% |
| Transmissibility: Scenario 3 | Peak in May 2024 | Scenario 3: Increase | $18,110 | -33.24% |
| Transmissibility: Scenario 4 | Peak in May 2024 | Scenario 4: Decrease | $39,181 | 44.44% |
| Transmissibility: Scenario 5 | Peak in May 2024 | Scenario 5: Peak in May 2024 and Peak in December 2024 | $636 | -97.66% |
| Residual VE | Assumes no new variant prior to March 2024. | Decrease the residual VE by 10% prior to March 2024 to simulate emergence of a new variant | $24,686 | -9.00% |
| Residual VE | Vaccine coverage<br>18-29 years = 4.3%,<br>30-49 years = 7.1%,<br>50-64 years = 20.0%,<br>65-74 years = 50.5%,<br>75+ years = 50.5% | Increase residual VE by increasing vaccine coverage in Sept 2023 to January 2024. Total coverage as follows:<br>18-29 years = 5.5%,<br>30-49 years = 9.1%,<br>50-64 years = 25.4%,<br>65-74 years = 76.0%,<br>75+ years = 76.0% | $36,180 | 33.38% |
| Vaccine Coverage (High Coverage, influenza) | Winter 2024 Vaccine<br>18-29 years = 3.3%,<br>30-49 years = 5.9%,<br>50-64 years = 17.6%,<br>65-74 years = 45.9%,<br>75+ years = 45.9%<br><br>Summer 2024 Vaccine | High Coverage scenario<br>Winter 2024 Vaccine<br>18-29 years = 4.2%,<br>30-49 years = 6.5%,<br>50-64 years = 40.0%,<br>65-74 years = 63.6%,<br>75+ years = 69.8% | $34,437 | 26.95% |

| Variable or assumption | Base-case value | Plausible alternative(s) or range of values | Incremental cost / QALY gained | Description of impact on ICER |
|---|---|---|---|---|
| | 18-29 years = 0%,<br>30-49 years = 0%,<br>50-64 years = 0%,<br>65-74 years = 20.1%,<br>75+ years = 20.1% | Summer 2024 Vaccine<br>18-29 years = 0%,<br>30-49 years = 0%,<br>50-64 years = 0%,<br>65-74 years = 20.1%,<br>75+ years = 69.8% | | |
| Vaccine Coverage (Ages 30 years and older) | Winter 2024 Vaccine<br>18-29 years = 3.3%,<br>30-49 years = 5.9%,<br>50-64 years = 17.6%,<br>65-74 years = 45.9%,<br>75+ years = 45.9%<br><br>Summer 2024 Vaccine<br>18-29 years = 0%,<br>30-49 years = 0%,<br>50-64 years = 0%,<br>65-74 years = 20.1%,<br>75+ years = 20.1% | Age 30 years and over scenario<br>18-29 years = 0%,<br>30-49 years = 5.9%,<br>50-64 years = 17.6%,<br>65-74 years = 45.9%,<br>75+ years = 45.9%<br><br>Summer 2024 Vaccine<br>18-29 years = 0%,<br>30-49 years = 0%,<br>50-64 years = 0%,<br>65-74 years = 20.1%,<br>75+ years = 20.1% | $29,325 | 8.11% |
| Vaccine Coverage (Ages 65 years and older) | Winter 2024 Vaccine<br>18-29 years = 3.3%,<br>30-49 years = 5.9%,<br>50-64 years = 17.6%,<br>65-74 years = 45.9%,<br>75+ years = 45.9%<br><br>Summer 2024 Vaccine<br>18-29 years = 0%,<br>30-49 years = 0%,<br>50-64 years = 0%,<br>65-74 years = 20.1%,<br>75+ years = 20.1% | Age 65 years and over scenario<br>18-29 years = 0%,<br>30-49 years = 0%,<br>50-64 years = 0%,<br>65-74 years = 45.9%,<br>75+ years = 45.9%<br><br>Summer 2024 Vaccine<br>18-29 years = 0%,<br>30-49 years = 0%,<br>50-64 years = 0%,<br>65-74 years = 20.1%,<br>75+ years = 20.1% | $51,390 | 89.45% |

| Variable or assumption | Base-case value | Plausible alternative(s) or range of values | Incremental cost / QALY gained | Description of impact on ICER |
|---|---|---|---|---|
| 2024 vaccine initial VE (95% CI LB) | against Infection: 57.1%, against Hospitalisation: 84.3% | against Infection: 30.6%, against Hospitalisation: 80.3% | $66,219 | 144.12% |
| 2024 vaccine initial VE (95% CI UB) | against Infection: 57.1%, against Hospitalisation: 84.3% | against Infection: 83.7%, against Hospitalisation: 87.5% | $13,156 | -51.50% |
| 2024 VE waning: Infection (95% CI LB) | Monthly waning: 4.8% | Monthly waning: 3.1% | $24,587 | -9.36% |
| 2024 VE waning: Infection (95% CI UB) | Monthly waning: 4.8% | Monthly waning: 6.8% | $30,643 | 12.96% |
| 2024 VE waning: Hospitalisation (95% CI LB) | Monthly waning: 1.4% | Monthly waning: 0.6% | $26,452 | -2.49% |
| 2024 VE waning: Hospitalisation (95% CI UB) | Monthly waning: 1.4% | Monthly waning: 2.4% | $27,989 | 3.18% |
| Proportion of infections considered symptomatic (-10%) | 0 - 14 years = 60.60%<br>15 - 19 years = 58.42%<br>20 - 29 years = 63.55%<br>30 - 39 years = 70.28%,<br>40 - 49 years = 72.64%,<br>50 - 59 years = 74.39%<br>60 - 64 years = 77.14%,<br>65 - 74 years = 80.98%,<br>75+ years = 86.73% | 0 - 14 years =54.54%<br>15 - 19 years = 52.58%<br>20 - 29 years = 57.20%<br>30 - 39 years = 63.25%<br>40 - 49 years = 65.38%<br>50 - 59 years = 66.95%<br>60 - 64 years = 69.42%<br>65 - 74 years = 72.88%<br>75+ years = 78.06% | $33,175 | 22.30% |
| Proportion of infections considered symptomatic (+10%) | 0 - 14 years = 60.60%<br>15 - 19 years = 58.42%<br>20 - 29 years = 63.55%<br>30 - 39 years = 70.28%,<br>40 - 49 years = 72.64%,<br>50 - 59 years = 74.39% | 0 - 14 years = 66.66%<br>15 - 19 years = 64.26%<br>20 - 29 years = 69.91%<br>30 - 39 years = 77.31%<br>40 - 49 years = 79.90%<br>50 - 59 years = 81.82%<br>60 - 64 years = 84.85%<br>65 - 74 years = 89.07% | $22,199 | -18.16% |

| Variable or assumption | Base-case value | Plausible alternative(s) or range of values | Incremental cost / QALY gained | Description of impact on ICER |
|---|---|---|---|---|
| | 60 - 64 years = 77.14%,<br>65 - 74 years = 80.98%,<br>75+ years = 86.73% | 75+ years = 95.40% | | |
| Hospitalisation Rates in the unvaccinated (-25%) | 0 - 14 years = 2.14%<br>15 - 19 years = 1.78%<br>20 - 29 years = 3.18%<br>30 - 39 years = 3.25%<br>40 - 49 years = 3.56%,<br>50 - 59 years = 4.52%<br>60 - 64 years = 9.65%<br>65 - 74 years = 15.68%<br>75+ years = 42.40% | 0 - 14 years = 1.61%<br>15 - 19 years = 1.34%<br>20 - 29 years = 2.39%<br>30 - 39 years = 2.44%<br>40 - 49 years = 2.67%,<br>50 - 59 years = 3.39%<br>60 - 64 years = 7.24%,<br>65 - 74 years = 11.76%,<br>75+ years = 31.80% | $39,129 | 44.25% |
| Hospitalisation Rates in the unvaccinated (<60 years of age, 33% of base case) | 0 - 14 years = 2.14%<br>15 - 19 years = 1.78%<br>20 - 29 years = 3.18%<br>30 - 39 years = 3.25%<br>40 - 49 years = 3.56%,<br>50 - 59 years = 4.52%<br>60 - 64 years = 9.65%<br>65 - 74 years = 15.68%<br>75+ years = 42.40% | 0 - 14 years = 0.71%<br>15 - 19 years = 0.59%<br>20 - 29 years = 1.05%<br>30 - 39 years = 1.07%<br>40 - 49 years = 1.17%<br>50 - 59 years = 1.49%<br>60 - 64 years = 9.65%<br>65 - 74 years = 15.68%<br>75+ years = 42.40% | $43,101 | 58.89% |
| Hospital mortality rate (-25%) | 0 - 14 years = 0.03%<br>15 - 19 years = 0.09%<br>20 - 29 years = 0.10%<br>30 - 39 years = 0.15%<br>40 - 49 years = 0.50%,<br>50 - 59 years = 1.19%<br>60 - 64 years = 1.57%,<br>65 - 74 years = 3.32%,<br>75+ years = 6.95% | 0 - 14 years = 0.02%<br>15 - 19 years = 0.07%<br>20 - 29 years = 0.07%<br>30 - 39 years = 0.11%<br>40 - 49 years = 0.37%<br>50 - 59 years = 0.89%<br>60 - 64 years = 1.18%<br>65 - 74 years = 2.49%<br>75+ years = 5.21% | $28,016 | 3.28% |
| Hospital mortality rate (+25%) | 0 - 14 years = 0.03%<br>15 - 19 years = 0.09% | 0 - 14 years = 0.04%<br>15 - 19 years = 0.11% | $26,293 | -3.07% |

| Variable or assumption | Base-case value | Plausible alternative(s) or range of values | Incremental cost / QALY gained | Description of impact on ICER |
|---|---|---|---|---|
| | 20 - 29 years = 0.10%<br>30 - 39 years = 0.15%<br>40 - 49 years = 0.50%,<br>50 - 59 years = 1.19%<br>60 - 64 years = 1.57%,<br>65 - 74 years = 3.32%,<br>75+ years = 6.95% | 20 - 29 years = 0.12%<br>30 - 39 years = 0.19%<br>40 - 49 years = 0.62%<br>50 - 59 years = 1.49%<br>60 - 64 years = 1.96%<br>65 - 74 years = 4.15%<br>75+ years = 8.68% | | |
| Hospital readmission rate (-25%) | 10.7% | 8.0% | $27,672 | 2.01% |
| Hospital readmission rate (+25%) | 10.7% | 13.3% | $26,581 | -2.01% |
| Post-discharge mortality rate (-25%) | No ICU or ventilator: 5.4%<br>ICU only: 10.40%<br>Ventilator: 10.40% | No ICU or ventilator: 4.08%<br>ICU only: 7.80%<br>Ventilator: 7.80% | $30,357 | 11.91% |
| Post-discharge mortality rate (+25%) | No ICU or ventilator: 5.4%<br>ICU only: 10.40%<br>Ventilator: 10.40% | No ICU or ventilator: 6.75%<br>ICU only: 13.00%<br>Ventilator: 13.00% | $24,517 | -9.62% |
| Post-discharge mortality rate (0%) | No ICU or ventilator: 5.4%<br>ICU only: 10.40%<br>Ventilator: 10.40% | No ICU or ventilator: 0.0%<br>ICU only: 0.0%<br>Ventilator: 0.0% | $47,231 | 74.12% |
| Rates of infection-related myocarditis (-25%) | 0 - 14 years = 0.122%<br>15 - 19 years = 0.095%<br>20 - 29 years = 0.078%<br>30 - 39 years = 0.067%<br>40 - 49 years = 0.093%,<br>50 - 59 years = 0.137%<br>60 - 64 years = 0.137%,<br>65 - 74 years = 0.160%,<br>75+ years = 0.208% | 0 - 14 years = 0.09%<br>15 - 19 years = 0.07%<br>20 - 29 years = 0.06%<br>30 - 39 years = 0.05%<br>40 - 49 years = 0.07%<br>50 - 59 years = 0.10%<br>60 - 64 years = 0.10%<br>65 - 74 years = 0.12%<br>75+ years = 0.16% | $27,214 | 0.32% |

| Variable or assumption | Base-case value | Plausible alternative(s) or range of values | Incremental cost / QALY gained | Description of impact on ICER |
|---|---|---|---|---|
| Rates of infection-related myocarditis (+25%) | 0 - 14 years = 0.122%<br>15 - 19 years = 0.095%<br>20 - 29 years = 0.078%<br>30 - 39 years = 0.067%<br>40 - 49 years = 0.093%,<br>50 - 59 years = 0.137%<br>60 - 64 years = 0.137%,<br>65 - 74 years = 0.160%,<br>75+ years = 0.208% | 0 - 14 years = 0.15%<br>15 - 19 years = 0.12%<br>20 - 29 years = 0.10<br>30 - 39 years = 0.08%<br>40 - 49 years = 0.12%<br>50 - 59 years = 0.17%<br>60 - 64 years = 0.17%<br>65 - 74 years = 0.20%<br>75+ years = 0.26% | $27,038 | -0.32% |
| Rates of long COVID (-25%) | Not hospitalised: 4.7%<br>Hospitalised: 9.6% | Not hospitalised: 3.5%<br>Hospitalised: 7.2% | $27,216 | 0.33% |
| Rates of long COVID (+25%) | Not hospitalised: 4.7%<br>Hospitalised: 9.6% | Not hospitalised: 5.9%<br>Hospitalised: 12.0% | $27,036 | -0.33% |
| Hospitalisation cost (-25%) | No ICU or ventilator: $28,693<br>ICU only: $42,750<br>Ventilator: $65,595 | No ICU or ventilator: $21,520<br>ICU only: $32,062<br>Ventilator: $49,196 | $32,897 | 21.28% |
| Hospitalisation cost (+25%) | No ICU or ventilator: $28,693<br>ICU only: $42,750<br>Ventilator: $65,595 | No ICU or ventilator: $35,866<br>ICU only: $53,437<br>Ventilator: $81,993 | $21,355 | -21.28% |
| Hospitalisation recovery cost (-25%) | $239.01 | $179.26 | $27,167 | 0.15% |
| Hospitalisation recovery cost (+25%) | $239.01 | $298.76 | $27,086 | -0.15% |
| Outpatient treatment cost (-25%) | $430.53 | $322.89 | $27,784 | 2.42% |
| Outpatient treatment cost (+25%) | $430.53 | $538.16 | $26,469 | -2.42% |
| Long COVID cost (-25%) | $581.60 | $436.20 | $27,216 | 0.33% |
| Long COVID cost (+25%) | $581.60 | $727.00 | $27,036 | -0.33% |

| Variable or assumption | Base-case value | Plausible alternative(s) or range of values | Incremental cost / QALY gained | Description of impact on ICER |
|---|---|---|---|---|
| Adverse event costs (-25%) | Any Grade 3 local adverse events: $30.83<br>Any Grade 3 systemic adverse events: $8.13<br>Anaphylaxis: $975.87<br>Myocarditis / Pericarditis: $12373.83 | Any Grade 3 local adverse events: $23.13<br>Any Grade 3 systemic adverse events: $6.10<br>Anaphylaxis: $731.90<br>Myocarditis / Pericarditis: $9,280.37 | $27,004 | -0.45% |
| Adverse event costs (+25%) | Any Grade 3 local adverse events: $30.83<br>Any Grade 3 systemic adverse events: $8.13<br>Anaphylaxis: $975.87<br>Myocarditis / Pericarditis: $12373.83 | Any Grade 3 local adverse events: $38.54<br>Any Grade 3 systemic adverse events: $10.17<br>Anaphylaxis: $1,219.84<br>Myocarditis / Pericarditis: $15,467.29 | $27,249 | 0.45% |
| Short-term infection period QALY loss (outpatient and hospitalisations) (-25%) | Not hospitalised: 0.003<br>Hospitalised general ward: 0.008<br>ICU: 0.016<br>ICU and ventilator: 0.03 | Not hospitalised: 0.0023<br>Hospitalised general ward: 0.006<br>ICU: 0.012<br>ICU and ventilator: 0.03 | $27,426 | 1.10% |
| Short-term infection period QALY loss (outpatient and hospitalisations) (+25%) | Not hospitalised: 0.003<br>Hospitalised general ward: 0.008<br>ICU: 0.016<br>ICU and ventilator: 0.03 | Not hospitalised: 0.0038<br>Hospitalised general ward: 0.010<br>ICU: 0.020<br>ICU and ventilator: 0.0375 | $26,833 | -1.08% |
| Adverse event QALY loss (-25%) | Grade 3 local: 0.00027<br>Grade 3 systemic: 0.00110<br>Anaphylaxis: 0.00192<br>Vaccine-induced myocarditis: 0.0019 | Grade 3 local: 0.00021<br>Grade 3 systemic: 0.00082<br>Anaphylaxis: 0.00144<br>Vaccine-induced myocarditis: 0.00143 | $26,979 | -0.54% |
| Adverse event QALY loss (+25%) | Grade 3 local: 0.00027<br>Grade 3 systemic: 0.00110<br>Anaphylaxis: 0.00192 | Grade 3 local: 0.00034<br>Grade 3 systemic: 0.00137<br>Anaphylaxis: 0.0024 | $27,276 | 1.10% |

| Variable or assumption | Base-case value | Plausible alternative(s) or range of values | Incremental cost / QALY gained | Description of impact on ICER |
|---|---|---|---|---|
| | Vaccine-induced myocarditis: 0.0019 | Vaccine-induced myocarditis: 0.00238 | | |
| Post-infection QALY loss (-25%) | Non-hospitalised during acute infection: 0.028<br>Hospitalised during acute infection: 0.122 | Non-hospitalised during acute infection: 0.0208<br>Hospitalised during acute infection: 0.0915 | $30,197 | 11.32% |
| Post-infection QALY loss (+25%) | Non-hospitalised during acute infection: 0.028<br>Hospitalised during acute infection: 0.122 | Non-hospitalised during acute infection: 0.0346<br>Hospitalised during acute infection: 0.1525 | $24,623 | -9.23% |
| Vaccine-related myocarditis rates: decrease | 0.0032% | 0.0002% | $27,120 | -0.024% |
| Vaccine-related myocarditis rates: increase | 0.0032% | 0.0035% | $27,127 | 0.003% |
| Perspective | Health care system | Societal | $15,765 | -41.88% |
| Discount rate | 5% | 0% | $16,978 | -37.41% |
| Discount rate | 5% | 1.5% | $20,571 | -24.17% |
| Discount rate | 5% | 3.5% | $24,609 | -9.28% |

QALY, quality-adjusted life-year; ICU, intensive care unit; CI, confidence interval; LB, lower bound; UB, upper bound

12. Ensemble model. Available at: https://covid19forecasthub.org/doc/ensemble/. at https://covid19forecasthub.org/doc/ensemble/.)

13. Covid-19 publications. Available at: https://www.healthdata.org/covid/publications. at https://www.healthdata.org/covid/publications.)

14. Coronavirus disease 2019 (COVID-19) epidemiology reports. 2023. (Accessed Oct 24, 2023, at https://www1.health.gov.au/internet/main/publishing.nsf/Content/covid-19_epidemiology_reports_australia_2022-23.htm.)

15. Coronavirus disease 2019 (COVID-19) epidemiology reports. 2021. (Accessed Oct 24, 2023, at https://www1.health.gov.au/internet/main/publishing.nsf/Content/covid-19_epidemiology_reports_australia_2021.htm.)

16. Admitted patient activity. 2023. (Accessed Oct 24, 2023, at https://www.aihw.gov.au/reports-data/myhospitals/intersection/activity/apc.)

17. Kohli MA, Maschio M, Lee A, et al. The potential clinical impact of implementing different COVID-19 boosters in fall 2022 in the United States. J Med Econ 2022;25:1127-39.

18. Gomez A, Kelly M, Sloan-Gardner T, Voo TV, Kirk M. Severity and Symptom Characteristics between Omicron and Delta SARS-CoV-2 Variant Infections in the Australian Capital Territory: A Cross-Sectional Study. Research Square; 2022.

25. Biddle N, Korda R. The experience of COVID-19 in Australia, including long-COVID–Evidence from the COVID-19 Impact Monitoring Survey Series, August 2022 ANU Centre for Social Research and. 2022.

32. Ham DI. Long-haulers and labor market outcomes: Federal Reserve Bank of Minneapolis; 2022.

33. Severity and Symptom Characteristics between Omicron and Delta SARS-CoV-2 Variant Infections in the Australian Capital Territory: A Cross-Sectional Study (Preprint from Research Square). 2022. (Accessed Oct 24, 2023, at https://doi.org/10.21203/rs.3.rs-1786210/v1 )

34. Demonstrating the utility of the COVID-19 Register. 2023. (Accessed Oct 24, 2023, at https://www.aihw.gov.au/reports/covid-19/demonstrating-the-utility-of-the-covid-19-register/contents/about.)

35. NSW Respiratory Surveillance Report - fortnight ending 18 November 2023. Epidemiological weeks 45 & 46, ending 18 November 20232023.
